# White Matter Cerebrovascular Reactivity: Effects of Microangiopathy and Proximal Occlusions on the Dynamic BOLD Response

**DOI:** 10.1101/2023.05.29.23290700

**Authors:** J Michael Gee, Xiuyuan Wang, Siddhant Dogra, Jelle Veraart, Koto Ishida, Seena Dehkharghani

## Abstract

**Introduction:** Cerebral microangiopathy often manifests as white matter hyperintensities (WMH) on T2-weighted MR images and is associated with elevated stroke risk. Large vessel steno-occlusive disease (SOD) is also independently associated with stroke risk, however, the interaction of microangiopathy and SOD is not well understood. Cerebrovascular reactivity (CVR) describes the capacity of cerebral circulation to adapt to changes in perfusion pressure and neurovascular demand, and its impairment portends future infarctions. CVR can be measured with blood oxygen level dependent (BOLD) imaging following acetazolamide stimulus (ACZ-BOLD). We studied CVR differences between WMH and normal-appearing white matter (NAWM) in patients with chronic SOD, hypothesizing additive influences upon CVR measured by novel, fully dynamic CVR maxima (*CVR_max_*).

**Methods:** A cross sectional study was conducted to measure per-voxel, per-TR maximal CVR (*CVR_max_*) using a custom computational pipeline in 23 subjects with angiographically-proven unilateral SOD. WMH and NAWM masks were applied to *CVR_max_*maps. White matter was subclassified with respect to the SOD-affected hemisphere, including: i. contralateral NAWM; ii. contralateral WMH iii. ipsilateral NAWM; iv. ipsilateral WMH. *CVR_max_* was compared between these groups with a Kruskal-Wallis test followed by a Dunn-Sidak post-hoc test for multiple comparisons.

**Results:** 19 subjects (age 50±12 years, 53% female) undergoing 25 examinations met criteria. WMH volume was asymmetric in 16/19 subjects with 13/16 exhibiting higher volumes ipsilateral to SOD. Pairwise comparisons of *CVR_max_*between groups was significant with ipsilateral WMH *CVR_max_* lower than contralateral NAWM (p=0.015) and contralateral WMH (p=0.003) when comparing in-subject medians and lower than all groups when comparing pooled voxelwise values across all subjects (p<0.0001). No significant relationship between WMH lesion size and *CVR_max_* was detected.

**Conclusion:** Our results suggest additive effects of microvascular and macrovascular disease upon white matter CVR, but with greater overall effects relating to macrovascular SOD than to apparent microangiopathy. Dynamic ACZ-BOLD presents a promising path towards a quantitative stroke risk imaging biomarker.

**BACKGROUND:** Cerebral white matter (WM) microangiopathy manifests as sporadic or sometimes confluent high intensity lesions in MR imaging with T2-weighting, and bears known associations with stroke, cognitive disability, depression and other neurological disorders^1–5^. Deep white matter is particularly susceptible to ischemic injury owing to the deprivation of collateral flow between penetrating arterial territories, and hence deep white matter hyperintensities (WMH) may portend future infarctions^6–8^. The pathophysiology of WMH is variable but commonly includes a cascade of microvascular lipohyalinosis and atherosclerosis together with impaired vascular endothelial and neurogliovascular integrity, leading to blood brain barrier dysfunction, interstitial fluid accumulation, and eventually tissue damage^9–14^.

Independent of the microcirculation, cervical and intracranial large vessel steno-occlusive disease (SOD) often results from atheromatous disease and is associated with increased risk of stroke owing to thromboembolic phenomena, hypoperfusion, or combinations thereof^15–17^. White matter disease is more common in the affected hemisphere of patients with asymmetric or unilateral SOD, producing both macroscopic WMH detectable by routine structural MRI, as well as microstructural changes and altered structural connectivity detected by advanced diffusion microstructural imaging^18, 19^. An improved understanding of the interaction of microvascular disease (i.e., WMH) and macrovascular steno-occlusion could better inform stroke risk stratification and guide treatment strategies when coexistent.

Cerebrovascular reactivity (CVR) is an autoregulatory adaptation characterized by the capacity of the cerebral circulation to respond to physiological or pharmacological vasodilatory stimuli^20–22^. CVR may be heterogeneous and varies across tissue type and pathological states^1, 16^. Alterations in CVR are associated with elevated stroke risk in SOD patients, although white matter CVR, and in particular the CVR profiles of WMH, are only sparsely studied and not fully understood^1, 23–26^. We have previously employed blood oxygen level dependent (BOLD) imaging following a hemodynamic stimulus with acetazolamide (ACZ) in order to measure CVR (i.e. ACZ-BOLD)^21, 27, 28^. Despite the emergence of ACZ-BOLD as a technique for clinical and experimental use, poor signal-to-noise characteristics of the BOLD effect have generally limited its interpretation to coarse, time-averaged assessment of the terminal ACZ response at arbitrarily prescribed delays following ACZ administration (e.g. 10-20 minutes)^29^. More recently, we have introduced a dedicated computational pipeline to overcome historically intractable signal-to-noise ratio (SNR) limitations of BOLD, enabling fully dynamic characterization of the cerebrovascular response, including identification of previously unreported, unsustained or transient CVR maxima (*CVR_max_*) following hemodynamic provocation^27, 30^.

In this study, we compared such dynamic interrogation of true CVR maxima between WMH and normal appearing white matter (NAWM) among patients with chronic, unilateral SOD in order to quantify their interaction and to assess the hypothesized additive effects of angiographically-evident macrovascular stenoses when intersecting microangiopathic WMH.

## METHODS

### Subject selection

ACZ-BOLD is obtained routinely in our practice as standard-of-care CVR estimations in patients with chronic SOD. Portions of the subject population have been previously studied in related investigations into the ACZ-BOLD technique^27, 30^. As reported previously, a population of 23 consecutive patients with angiographically-proven, unilateral, chronic SOD undergoing ACZ-BOLD between May 8, 2017-October 10, 2020 for recurrent transient ischemic attack or minor stroke were evaluated using a cross-sectional study design with Institutional Review Board approval. The STROBE cross-sectional reporting guidelines were used to inform the study design and in preparation of the manuscript^31^.

Subjects were excluded if there was evidence for prior surgery, prior infarctions larger than lacunae, hemorrhage, or major anatomical deformities on MRI images. Inclusion criteria included catheter angiographic evidence of unilateral SOD and successful completion of the study protocol.

### MR Imaging

Subjects were scanned on a 3T whole-body system (Prisma, Siemens Healthineers, Erlangen, Germany) with a 64-channel head coil as previously described^27, 30^. The protocol included: i. 1 mm isotropic T1-weighted MPRAGE (TR=2300 ms, TE=2.9 ms, flip angle=9°, voxel size=1×1×1 mm^3^); ii. 3D isotropic FLAIR TR 6 s, TE 325 ms, flip angle 120°, voxel size 1×1×1 mm^3^; iii. 20-minute continuous BOLD imaging performed with a gradient-echo EPI sequence (TR=2000ms, TE=40 ms, 1 NEX, flip angle=90°, voxel size=2×2×5 mm^3^, whole-brain coverage); iv. Gradient-echo magnitude and phase field maps to correct for spatial distortions in BOLD images.

#### ACZ-BOLD paradigm

ACZ (1 gram dissolved in 10 mL saline) was infused intravenously over 3-5 minutes following an initial 4–5-minute baseline BOLD, without interruption of BOLD imaging upon or during injection. The total number of BOLD volumes and repetition times, and thus the effective post-BOLD duration, varied slightly between studies to ensure whole brain coverage, but exceeded 20 minutes in all cases.

#### Image processing

A custom computational pipeline developed by our group was employed for multifold enhancement in SNR characteristics of the ACZ-BOLD response, as elaborated previously^27, 30^; briefly, a Marchenko-Pastur Principle Component Analysis (MP-PCA)-based denoising algorithm was developed to identify spatially-varying and non-signal bearing noise sources in a local eigenspectrum permitting nulling of thermal noise contributions while preserving the dynamic time-signal course (TSC)^21, 27, 30^. Denoised BOLD images and field maps were processed in FSL Melodic for motion correction, unwarping, slice-timing correction, and spatial smoothing (FMRIB Software Library). Initial and terminal 30 frames were discarded due to low-pass filtering-related ringing artifact.

In order to capture the full range of CVR dynamics, including unsustained and transient or occult maxima in the cerebrovascular response, maximal CVR (*CVR_max_*) was isolated in lieu of conventional measures of terminal CVR (*CVR_end_*) as detailed previously^27^. The maximal CVR per-voxel, per-TR, across the TSC was computed from denoised BOLD data as expounded in initial reports^27, 30^

#### Mask generation

WMH masks were generated by manual segmentation of discrete supratentorial WMH on isotropic 1×1×1 mm^3^ FLAIR images using the 3D Slicer Segment Editor module^32, 33^ (Figure 1A and 1D). Segmentation was performed by a neuroradiology fellow (JMG), blinded to knowledge of the hemisphere affected by SOD, under the supervision of a subspecialty certified neuroradiologist (SDe) with greater than 10 years of experience in cerebrovascular imaging and advanced image processing. Hemispheric total WM and gray matter (GM) masks were generated in Freesurfer v7.1.1 from T1-weighted images^34^ (Figure 1B) and corresponding total WM volumes were recorded per-hemisphere, per subject. Real-time evaluation of the full multiplanar 3D FLAIR data set with manual reformation was permitted in order to facilitate WMH identification and segmentation, while also improving sensitivity and specificity for affected areas near the ventricular and cortical surface. Manual magnification and window-level operations were permitted to further improve conspicuity and confidence in lesion identification and segmentation. NAWM masks were constructed by subtraction of the WMH mask from the total WM mask (Figure 1C). GM masks were dilated by 4 voxels and subtracted from WMH and NAWM masks in order to limit contamination of WM by cortical BOLD signal while minimizing cropping of juxtacortical WMH lesions. Confluent areas of white matter disease were circumscribed to include contiguously affected areas. Individual WMH lesion volumes (in cc) were recorded, and total volume of WMH in each hemisphere was computed as the total sum of all individual lesion volumes.

**Figure 1.**
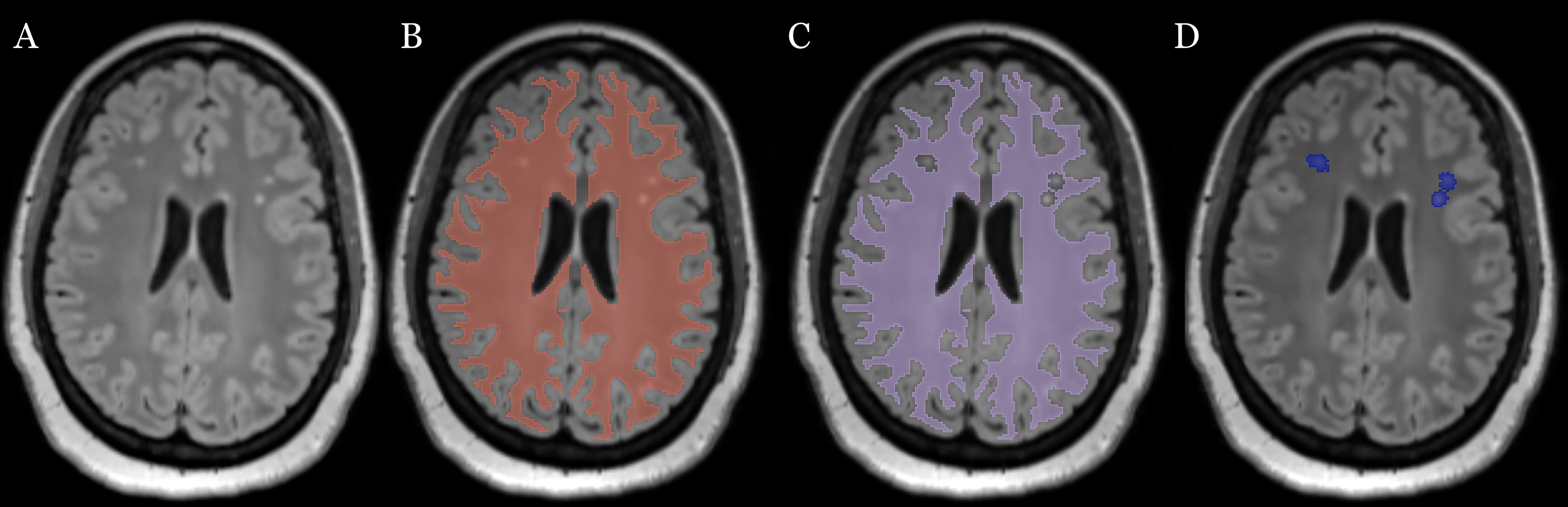
White matter mask generation. (A) FLAIR image demonstrating multiple white matter hyperintensities (WMH) in the bilateral frontal lobes in a patient with angiographically documented left-sided steno-occlusive disease. (B) Cerebral hemisphere white matter masks generated from 3D T1 volumes. (C) Normal-appearing white matter (NAWM) masks generated by subtracting the lesion mask from the cerebral white matter mask. (D) Manual WMH segmentation and masking.

#### White matter classification and volumes

Voxel-wise *CVR_max_* maps were calculated for both cerebral hemispheres. Total WM, NAWM and WMH masks were applied to *CVR_max_* images (Figure 2) with custom MATLAB scripts (version R2022b, The MathWorks, Natick, MA). Due to global differences in *CVR_max_* values between subjects, *CVR_max_* were scaled to a median of 0 and interquartile range (IQR) of 1 using the formula [*CVR*_vox_ – median(*CVR_all_*)]/IQR(*CVR_all_*) where *CVR_vox_* represents the single-voxel *CVR_max_* and *CVR_all_* represents *CVR_max_* values of all WM voxels in both hemispheres.

**Figure 2.**
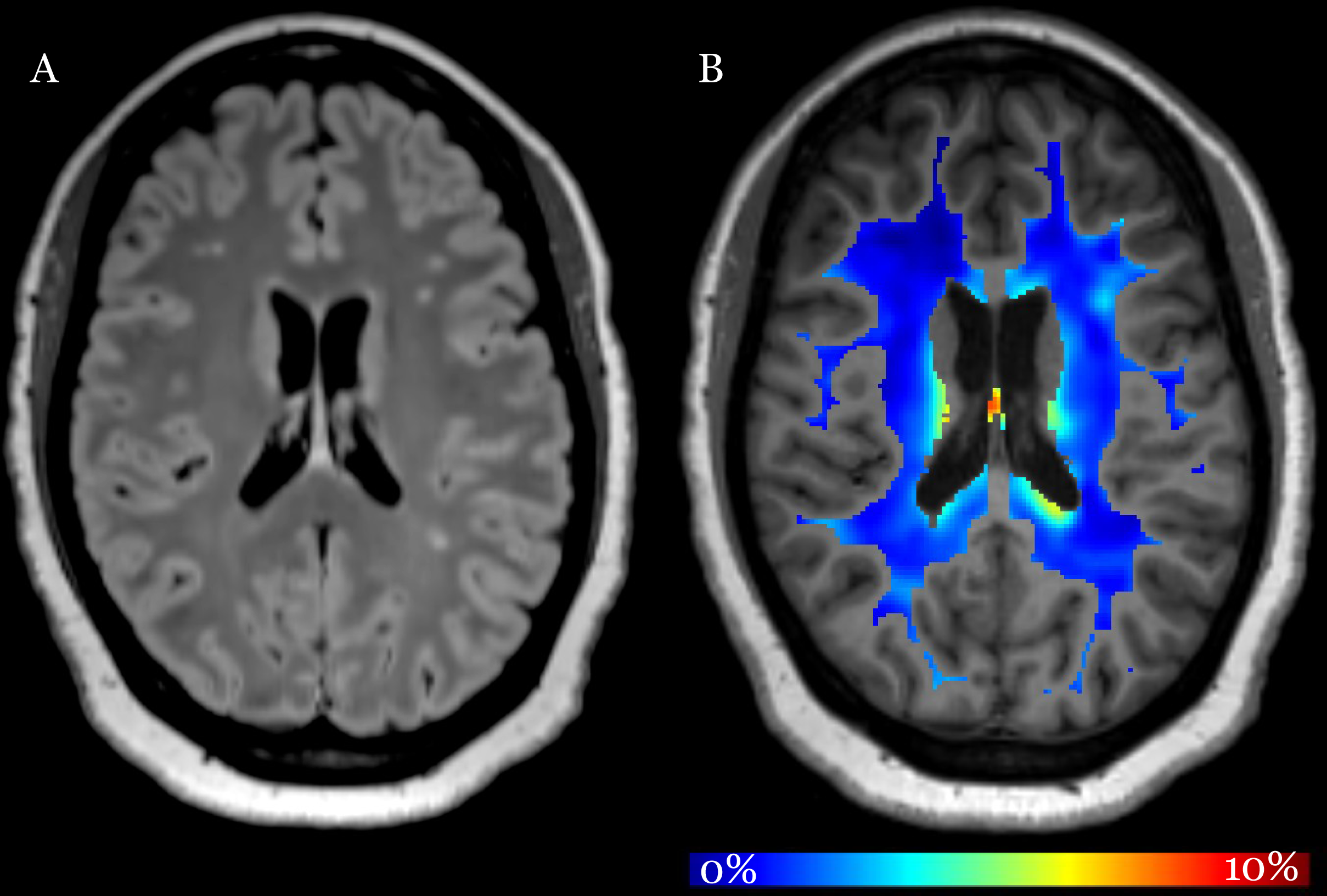
(A) FLAIR image demonstrating multiple white matter hyperintensities in the cerebral white matter bilaterally in a patient with left-sided steno-occlusive disease. (B) Total white matter mask applied to the *CVR_max_* map superimposed on a T1-weighted image. *CVR_max_* is expressed as a percent-change in BOLD signal following acetazolamide administration using the ACZ-BOLD paradigm. Dark blue voxels correspond to relatively low *CVR_max_*values and green/red voxels to high *CVR_max_* values.

WMH volumes were compared across hemispheres with respect to the side of the proximal arterial steno-occlusive lesions. A WMH asymmetry index was established to facilitate investigation of CVR differences between hemispheres relative to their respective burden of WMH. The presence of a meaningful difference in WMH volumes was set empirically at a 10% fractional difference in volumes, representing a conservative estimation based in prior meta-analysis documenting approximately 10% differences in WM diffusional change in hemispheres affected by unilateral anterior circulation steno-occlusion^18^. A 10% threshold was thus anticipated to constitute a conservative initial approximation likely to capture subtle asymmetries in WM lesion burden. The fractional difference in WMH volume between hemispheres was computed as: |(*Vol_ipsi_*-*Vol_contra_*)/(*Vol_ipsi_*+*Vol_contra_*)| > 0.1.

All WM voxels in every subject were thereby classified into one of four tissue groups based in the above procedure, with respect to FLAIR imaging findings (NAWM vs. WMH) and laterality of SOD. The hemisphere affected by SOD was denoted by subscripted *ipsi* for WMH ipsilateral to macrovascular steno-occlusion, and subscripted *contra* for the contralateral hemisphere. White matter classes were thus grouped as: *NAWM_contra_, WMH_contra_, NAWM_ipsi_,* and *WMH_ipsi_*.

### Statistical Analysis

Statistical analysis was performed in MATLAB. A Lillefors test was used to assess for normal distributions within each group. A Student’s T-test was used to compare WM group volumes between hemispheres. Median *CVR_max_* values in each tissue group were compared within and between subjects by the Kruskal-Wallis rank sum test followed by a Dunn-Sidak post-hoc multiple pairwise comparisons test. A similar approach was taken to compare pooled voxel-wise data across all subjects. Summary *CVR_max_* data across subjects is reported as median[IQR]. To explore the relationship between CVR and WMH lesion volume or total hemispheric WMH burden, a Pearson correlation coefficient (*r*) was calculated. Statistical significance was set at alpha=0.05.

## RESULTS

A total of 19 subjects undergoing 25 examinations met criteria. The average age of subjects was 50±12 years (range 27-74 years) with 10/19 (53%) subjects female. Detailed demographic features of the study population were reported previously^27^. One subject was excluded due to extensive burden of remote infarctions with encephalomalacia and gliosis. Three subjects were excluded due to absence of a 3D-FLAIR image.

### Comparison of total WM volume and WMH volume by hemisphere

Interhemispheric WMH volume was asymmetric in 16/19 subjects based on the WMH asymmetry threshold of 10%. NAWM volume in the ipsilateral hemisphere was 212.7.7±27.5 cc and 219.0±27.4 cc in the contralateral hemisphere (95% CI [2.3,10.4], p=0.004). WMH volume in the ipsilateral hemisphere was 2.9±3.0 cc and 1.5±1.7 cc in the contralateral hemisphere (95% CI [-2.3,-0.4], p=0.006). In 13/16 subjects with asymmetric hemispheric WMH volume, the hemisphere affected by SOD also demonstrated the asymmetrically higher WMH volume.

### WM CVR Profiles Relative to Hemispheric SOD

Comparison of median CVR in *WMH_ipsi_* to *WMH_contra_* within subjects revealed *CVR_max_* profiles that were lower in *WMH_ipsi_* in 19/25 studies. For 5/6 studies in whom *CVR_max_* was not lower in *WMH_ipsi_*, there was a more generalized hemispheric reduction in CVR in the contralateral hemisphere, consistent with counter-steal phenomena reported in earlier studies (see below).

Median[IQR] tissue class *CVR_max_* was compared across all subjects (Figure 3A). Median *CVR_max_* values of each tissue group across subjects were *NAWM_contra_* 0.11[0.41], *WMH_contra_* 0.17[0.66], *NAWM_ipsi_* -0.12[0.51], and WMH_ipsi_ -0.34[0.67] with median *CVR_max_*of *WMH_ipsi_* significantly lower than that of *WMH_contra_* (p=0.003) and *NAWM_contra_* (p=0.015).

**Figure 3.**
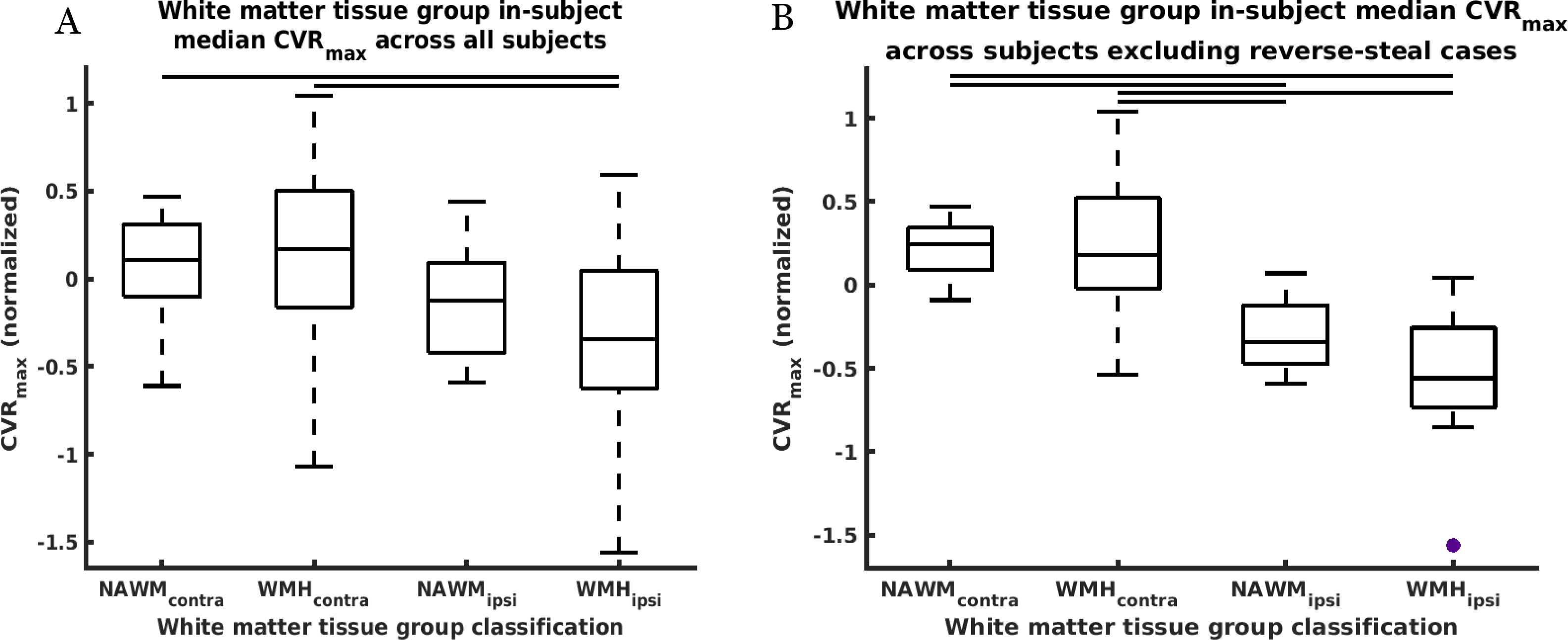
(A) Groupwise white matter tissue group *CVR_max_* boxplot summaries across all subjects. Median *CVR_max_* values for each tissue class were derived from the per-subject median of in-subject median tissue class *CVR_max_* values. Median *CVR_max_* in *WMH_ipsi_* was significantly lower than *NAWM_contra_* or *WMH_contra_*. (B) Groupwise white matter tissue group *CVR_max_* boxplot summaries across subjects with removal of subjects demonstrating reverse-steal (*Robinhood effect*). Median *CVR_max_* in *WMH_ipsi_*was significantly lower than *NAWM_contra_* or *WMH_contra_*. *NAWM_ipsi_* was lower than both *NAWM_contra_* and *WMH_contra_*, further supporting the idea that macrovascular disease contributes more to impaired CVR than microvascular disease. The central line of the boxplots represent the median; box edges represent 25th and 75th percentiles; whiskers show extreme datapoints (1.5*IQR from the median). Outlier values greater than 1.5*IQR from the median are shown with a violet circle. Black horizontal bars represent statistical significance (p<0.05).

Results of between-subject group analysis using pooled voxels for comparison between tissue classes yielded median[IQR] *CVR_max_* for *NAWM_contra_* 0.12[0.93], *WMH_contra_* 0.11[0.92], *NAWM_ipsi_* -0.13[0.99], and *WMH_ipsi_* -0.20[0.83]. Pairwise comparisons between all groups were significant with p<0.0001. When all WM voxels from all subjects were pooled and compared, *WMH_ipsi_ CVR_max_* values were significantly lower than all other groups and demonstrated a leftward cumulative distribution curve skew (Figure 4A; p<0.0001), suggesting additive effects of microvascular and macrovascular disease in CVR estimation.

**Figure 4.**
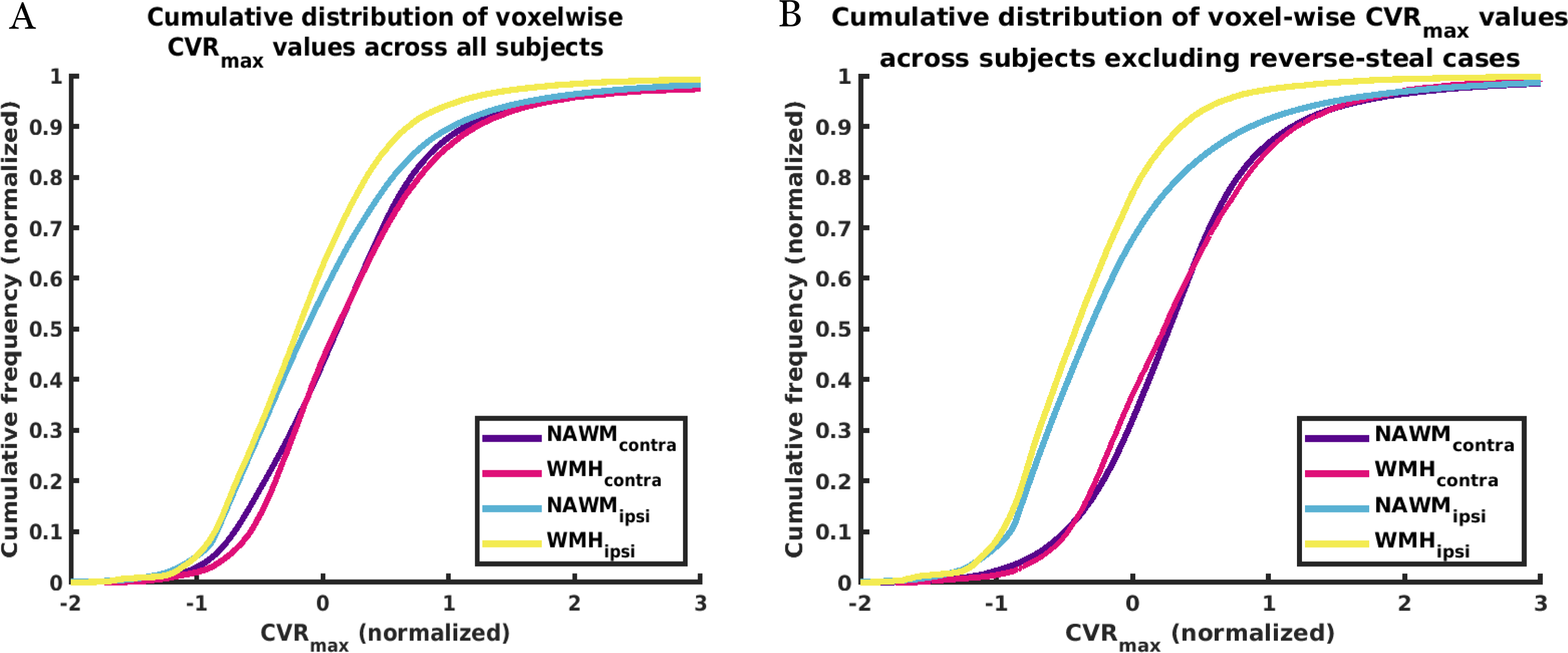
(A) Voxel-wise cumulative distribution function (CDF) curves of pooled *CVR_max_* values from all subjects. *WMH_ipsi_* median *CVR_max_* was lowest and CDF curves were most leftward skewed (yellow). *NAWM_contra_*median *CVR_max_* values were highest and CDF curves were most rightward skewed (blue). Median *CVR_max_* of both ipsilateral hemisphere groups were lower than both contralateral hemisphere groups. Median *CVR_max_* values and CDF curves in *NAWM_contra_* (violet) and *WMH_contra_* (magenta) were very similar and small differences are of questionable physiological significance. All pairwise comparison differences were statistically significant (p<0.0001). (B) Voxel-wise cumulative distribution curves of pooled *CVR_max_* values with studies demonstrating reverse-steal (*Robinhood effect*) removed. There was increased leftward skew of the *WMH_ipsi_* CDF relative to the other tissue classes, highlighting potential additive effects of microvascular disease to impaired CVR.

### Comparative Influence of Macrovascular disease vs. Microvascular Disease on CVR

In order to test for differences in CVR in relation to microvascular disease, within-hemisphere WMH and NAWM median CVR across all subjects were compared (Figure 3A). In 16/25 studies, median *CVR_max_* in *WMH_ipsi_* was lower than *NAWM_ipsi_*. Higher median *CVR_max_* values in *NAWM_ipsi_* relative to *WMH_ipsi_* were not statistically different (p=0.86). Similarly, higher *CVR_max_* values in *WMH_contra_* relative to *NAWM_contra_* were not statistically different (p=0.99).

When comparing pooled voxels across all subjects in the ipsilateral hemisphere, median *CVR_max_* values were lower in *WMH_ipsi_* than *NAWM_ipsi_* (p<0.0001), however, within the contralateral hemisphere, *WMH_contra_* and *NAWM_contra_* median *CVR_max_* values were 0.11[0.93] vs 0.12[0.92] and their distributions appeared nearly identical (Figure 4A).

### WM CVR Profiles in subjects with counter steal profiles

We have previously reported the occasional presence of paradoxical counter-steal (i.e. *Robinhood*) profiles among patients with large interhemispheric communicating arteries whereby a generalized reversal of CVR is noted between the diseased and normal hemispheres^27, 30^. In the current study, 9 such cases were encountered, demonstrating a quantitative and qualitative relative global reduction of *CVR_max_*in the hemisphere contralateral to the SOD hemisphere. In view of its presence, additional post-hoc analysis was undertaken following exclusion of the small number of such cases. With these subjects removed, median *CVR_max_*values in *NAWM_contra_* were 0.24[0.25], *WMH_contra_*0.18[0.54], *NAWM_ipsi_* -0.34[0.35] and *WMH_ipsi_*-0.56[0.48] (Figure 3B).

Pairwise comparisons demonstrated statistically lower *CVR_max_* in *WMH_ipsi_* compared to *NAWM_contra_* (p<0.0001) and *WMH_contra_* (p<0.0001). *NAWM_ipsi_ CVR_max_* was lower than *NAWM_contra_* (p =0.0006). *NAWM_ipsi_ CVR_max_* was lower than *WMH_contra_* (p=0.0019), suggesting a potentially larger contribution to decreased *CVR_max_*from SOD compared to apparent microangiopathy. Similarly, higher *CVR_max_*in *NAWM_ipsi_* compared to *WMH_ipsi_* were not statistically significant (p=0.78) and higher *CVR_max_* values in *NAWM_contra_* compared to *WMH_contra_* did not reach statistical significance (p=0.99).

Median[IQR] pooled voxel-wise *CVR_max_* values were *NAWM_contra_* 0.27[0.78], WMH_contra_ 0.23[0.91], *NAWM_ipsi_* -0.32[0.87] and *WMH_ipsi_* -0.41[0.72]. Pairwise comparisons between all groups were statistically significant (p<0.0001). When compared to the whole group analysis, there was an increased relative leftward skew of the *WMH_ipsi_* values and rightward skew of the *NAWM_contra_* values (Figure 4B).

### Relationship between WMH lesion burden and CVR

Potential interactions of lesion size to CVR in both hemispheres were examined. In a preliminary investigation, we considered the relationship between individual WMH size and CVR signatures. However, initial analysis revealed marked clustering of WMH volume from across the entire population to a relatively narrow overall range spanning less than 100 mm^3^, across which many-fold differences in *CVR_max_* were observed. There was no significant correlation of lesion size and *CVR_max_* (contralateral hemisphere r=0.05, p= 0.4; ipsilateral hemisphere r=-0.02, p=0.76). In contralateral hemispheres, the slope of lesion size versus *CVR_max_* was -0.00004 and in ipsilateral hemispheres was 0.00013. Here again, because the relative contribution of WMH appeared much smaller than the presence of SOD, further evaluation of this interaction was not studied. It was therefore elected to explore the relationship between total hemispheric WMH volume and CVR signatures in the respective hemispheres on the basis of observed worsening of CVR in hemispheres more heavily burdened by SOD. Total hemispheric WMH burden was compared to the median *CVR_max_*. There was, however, no significant correlation between WMH burden and *CVR_max_* in the ipsilateral hemisphere (r=0.19, p=0.37) or the contralateral hemisphere (r=-0.07, p=0.74).

## DISCUSSION

In this study, we used ACZ-BOLD estimations of CVR to study the association and interactions of microvascular and macrovascular disease to white matter CVR in a population of patients with angiographically documented unilateral SOD. While a far stronger influence upon CVR by SOD is suggested by the results, a potentially strong additive effect of microvascular disease upon CVR impairment is also apparent, with *CVR_max_*of WMH ipsilateral to large vessel SOD lower than that of all other WM tissue classes.

The presence of microvascular disease did not emerge as a strong predictor of CVR impairment within hemispheres, although its contribution again appears disproportionately to impact hemispheres burdened by macrovascular SOD. As expected, there was a significant contribution of macrovascular disease in the ipsilateral hemisphere to impaired CVR, outweighing any effect of microvascular disease in the contralateral hemisphere. Within our study population, proximal large vessel macrovascular disease was more strongly associated with CVR impairment than local microangiopathic changes, irrespective of individual WMH characteristics or total burden. It bears mention that while lesion size represents the only measurable predictor of severity for any given WMH in our study, it may be a poor predictor of perfusional impairment; the often small size of WMH precludes reliable estimation of blood flow in individual lesions using conventional techniques, which we anticipated would impart unrecoverable bias in favor of smaller WMH, hence our foregoing such an approach, and instead focusing on lesion size as a reasonable heuristic using isotropical high-resolution imaging. Further, the degree of signal abnormality for individual WMH is not readily quantifiable using conventional image weighting, although emergent fingerprinting techniques as used in past studies present potential opportunities to more rigorously quantify the tissue changes, irrespective of lesion size^35, 36^. Notwithstanding, the intersection of the macrovascular and microvascular changes in our study seems, unsurprisingly, to heighten the likelihood for CVR reductions, although the long-term prognostic significance of such CVR signatures remains to be established. While hemispheric WMH volume was more commonly higher in the hemisphere affected by SOD, there were examples where the opposite was true. Among these patients, the relationship between CVR in WMH and SOD remains indeterminate.

Robust automated algorithms for WMH segmentation are limited, although a few hold promise^37, 38^. Generation of WMH masks is a difficult and laborious task and intrinsically subjective. When manually segmenting WMH, the borders of WMH may be difficult to discern due to volume averaging and uncertainty arising from variable flip angles induced across the brain. These limitations can severely affect automated segmentation, and may introduce errors, favoring manual segmentation as performed herein^39^.

We hasten to add that NAWM is not equivalent to normal WM. Multiple studies, primarily within the multiple sclerosis literature, have demonstrated abnormalities of NAWM in the vicinity of WMH using various measures and underscoring the imperfect sensitivity of this approach despite its universal clinical adoption^40–42^. In our study, there were subsets of NAWM voxels that demonstrated very low CVR. NAWM was defined as all WM that was neither segmented in the WMH mask nor subtracted by the dilated gray matter masks. The potential influence of the post-processing pipeline merits dedicated analysis together with investigation of CVR in subsets of NAWM based on anatomical location, vascular territories, and geographical and morphological features of WMH.

Similarly, while WMH was used to identify regions of microangiopathy, a wide range of other pathologies may result in its development on fluid-sensitive brain MRI pulse sequences. Importantly, WM pathology related to small vessel disease may, itself, differ histologically depending on the nature of the disease. It is recognized that in early small vessel disease WMH may represent decreased mobility of interstitial fluid, whereas more advanced appearing WMH may represent gliosis, with an indeterminate effect upon measured CVR. The ability, even for expert readers using isotropic 3D imaging at high field (3T and above), to discriminate the two on structural brain MRI remains subjective, and here as well future studies may benefit from emergent quantitative imaging techniques to improve characterization of white matter pathology^36^.

Several additional study limitations merit discussion, including its retrospective nature, which could have imparted bias in patient selection. A consecutive population of patients presenting in our routine clinical cerebrovascular imaging practice were studied, with no a priori assumption that significant CVR impairment would be present. The relatively small study population could have reduced our statistical power to identify subtle interactions between study variables, although this represents the largest such study using ACZ-BOLD for characterization of WM CVR signatures, and to date only the third report of dynamic CVR measurement for extraction of CVR maxima from the pharmacologic stimulus. The spatial resolution of our BOLD imaging protocol (voxel size = 2 x 2 x 5 mm^3^) may have limited evaluation of CVR changes in small WMH, causing partial volume averaging. Steps were taken to minimize contamination from gray matter CVR by subtracting dilated gray matter masks from the white matter masks; however, juxtacortical WMH could have been subjected to mild morphologic and therefore quantitative erosion. Importantly, juxtacortical lesions were far less commonly observed than periventricular and subcortical WMH in our population, and any effects of unintended erosion is anticipated to have been small. Ultrahigh field MRI may provide a valuable path to improved spatial resolution and more localized BOLD effects, but until recently was unapproved for non-experimental clinical use^43^. Additionally, as discussed above, while coincident macrovascular and microvascular ischemic risks appear to portend greater impairment in CVR, the prognostic significance has yet to be established in a prospective or longitudinal cohort.

Nevertheless, our results suggest interesting interactions of microvascular and macrovascular disease that may inform stroke risk. Dynamic BOLD-CVR presents a promising path forward to quantitative prognostic biomarker development, and future studies will focus on further delineation of CVR signature in SOD patients.

## Data Availability

Data may be made available upon reasonable request.

## ACKNOWLEDGMENTS

None

